# Pharmacogenetics of tuberculosis treatment toxicity and effectiveness in a large Brazilian cohort

**DOI:** 10.1101/2023.08.30.23294860

**Authors:** Gustavo Amorim, James Jaworski, Marcelo Cordeiro-Santos, Afrânio L. Kritski, Marina C. Figueiredo, Megan Turner, Bruno B. Andrade, Digna R. Velez Edwards, Adalberto R. Santos, Valeria C. Rolla, Timothy R. Sterling, David W. Haas, the Regional Prospective Observational Research in Tuberculosis (RePORT)-Brazil network

## Abstract

**Background:** Genetic polymorphisms have been associated with risk of anti-tuberculosis treatment toxicity. We characterized associations with adverse events and treatment failure/recurrence among adults treated for tuberculosis in Brazil.

**Methods:** Participants were followed in Regional Prospective Observational Research in Tuberculosis (RePORT)-Brazil. We included persons with culture-confirmed drug-susceptible pulmonary tuberculosis who started treatment between 2015-2019, and who were evaluable for pharmacogenetics. Treatment included 2 months of isoniazid, rifampin or rifabutin, pyrazinamide, and ethambutol, then 4 months of isoniazid and rifampin or rifabutin, with 24 month follow-up. Analyses included 43 polymorphisms in 20 genes related to anti-tuberculosis drug hepatotoxicity or pharmacokinetics. Whole exome sequencing was done in a case-control toxicity subset.

**Results:** Among 903 participants in multivariable genetic association analyses, *NAT2* slow acetylator status was associated with increased risk of treatment-related grade 2 or greater adverse events, including hepatotoxicity. Treatment failure/recurrence was more likely among *NAT2* rapid acetylators, but not statistically significant at the 5% level. A *GSTM1* polymorphism (rs412543) was associated with increased risk of treatment-related adverse events, including hepatotoxicity. *SLCO1B1* polymorphisms were associated with increased risk of treatment- related hepatoxicity and treatment failure/recurrence. Polymorphisms in *NR1/2* were associated with decreased risk of adverse events and increased risk of failure/recurrence. In whole exome sequencing, hepatotoxicity was associated with a polymorphism in *VTI1A*, and the genes *METTL17* and *PRSS57*, but none achieved genome-wide significance.

**Conclusions:** In a clinical cohort representing three regions of Brazil, *NAT2* acetylator status was associated with risk for treatment-related adverse events. Additional significant polymorphisms merit investigation in larger study populations.

## Background

Tuberculosis is a leading cause of death from a single infectious agent worldwide.[1] Standard treatment for drug-sensitive tuberculosis involves two months of isoniazid, rifampin, pyrazinamide, and ethambutol, then four months of isoniazid and rifampin.[2, 3] Among patients who adhere to tuberculosis treatment in clinical trials, cure rates are as high as 97%.[4] However, in real-world settings only an estimated 86% have successful outcomes.[1] Factors that limit success include adverse drug reactions (ADR), acquired drug resistance during treatment, premature treatment discontinuation, and loss to follow-up.[5] Hepatotoxicity is the most frequent ADR with tuberculosis treatment, with reported rates from 2% to 28%,[6] with even higher rates among people with HIV (PWH).[7–9].

Treatment failure affects fewer than 5% of patients.[1, 2] While tuberculosis treatment is considered to be effective among PWH, death during treatment and loss to follow-up may be more frequent.[1, 10–12] Another concern is tuberculosis recurrence, whereby patients who complete treatment subsequently develop tuberculosis due to relapse or exogenous reinfection. Following standard therapy, approximately 4% of patients experience relapse or require retreatment within two years.[13, 14] Recurrence is reported to be more likely in PWH, particularly due to reinfection, and especially before availability of effective antiretroviral therapy (ART).[15–17]

Isoniazid is metabolized by N-acetyltransferase 2 [18], and *NAT2* loss-of-function alleles are frequent [18–20]. One or two copies of such alleles confer intermediate or slow acetylator phenotypes, respectively, and progressively greater isoniazid exposure [18–20]. Slow acetylator alleles have been reported to increase the risk for isoniazid hepatotoxicity [21–23].

Polymorphisms in additional genes have been reported to affect isoniazid and other anti-tuberculosis drugs, including *SLCO1B1* with rifampin [24], and *GSTM1* and *CYP2E1* with isoniazid [25], although less well replicated than for *NAT2*.

Improved understanding of relationships between human genetics and tuberculosis treatment toxicity and effectiveness could inform treatment strategies, which might include point-of-care pre-treatment genotyping. We characterized associations between selected genetic variants, treatment toxicity and effectiveness in a large, prospective cohort study in Brazil.

## Methods

### Study design and population

Participants were from Regional Prospective Observational Research in Tuberculosis (RePORT) Brazil, a cohort study of individuals with newly diagnosed, culture-confirmed, pulmonary tuberculosis. Between June 2015 and June 2019, participants were enrolled at five sites across three regions in Brazil - Rio de Janeiro, Salvador, and Manaus. RePORT-Brazil is broadly representative of tuberculosis cases in Brazil.[26] For this study, we included RePORT-Brazil participants with drug-susceptible tuberculosis on standard tuberculosis therapy, all of whom provided samples for genetics. Participants were followed for 24 months to assess treatment response and recurrence. RePORT-Brazil excluded individuals who previously received anti-tuberculosis therapy for ≥7 days, received >7 days of fluoroquinolone therapy within 30 days prior to enrollment, were pregnant or breastfeeding, or planned to leave the region during follow-up.

### Data collection and definitions

Clinical, demographic, laboratory (including sputum mycobacterial culture), and outcome data were collected at four study visits per RePORT-Brazil protocol: at treatment initiation (baseline), month 1 and month 2 after treatment initiation, and at end of treatment, which was typically 6 months after initiation. Standard therapy included isoniazid, rifampicin or rifabutin, pyrazinamide, and ethambutol for two months, followed by isoniazid and rifampicin or rifabutin for four months. Telephone follow-up occurred every 6 months thereafter until 24 months.

Participants were tested for HIV at baseline unless already known to be positive. Diabetes status at baseline was based on self-report. Directly observed therapy (DOT) was prescribed at treatment initiation.

Outcomes of interest included tuberculosis treatment toxicity and effectiveness (i.e., treatment failure and recurrence). Treatment failure was defined as remaining sputum culture-positive or smear-positive at month 5 or later during treatment. Recurrence was defined as culture-confirmed tuberculosis or symptoms consistent with tuberculosis after apparent cure or treatment completion. Primary toxicity outcomes were any grade ≥2 ADR that was considered at least possibly related to tuberculosis treatment, and grade ≥2 hepatotoxicity that was considered at least possibly related to tuberculosis treatment. We also considered grade ≥3 ADR related to TB treatment. Categories of physician-assigned attribution of causality were “possibly”, “likely”, or “definitely” related to tuberculosis treatment. Secondary analyses considered any adverse event.

Cases selected for whole exome sequencing were defined as having grade ≥3 treatment-related toxicity, permanent drug discontinuation due to treatment toxicity, or grade ≥2 treatment-related hepatotoxicity. Controls were matched on disease risk score, which was derived from age, sex, self-reported race, HIV status, *NAT2* acetylator group, and *CYP2B6* metabolizer group (if on efavirenz). Models are adjusted for sex, age, and 4 PCs.

### Genetic Polymorphisms

Human DNA was extracted from whole blood. We genotyped 45 selected polymorphisms in 21 genes relevant to anti-tuberculosis drug-induced hepatotoxicity (*ABCB1, ABCC2, AGBL4, BACH1, CUX2, CYP2E1, CYP7A1, GSTM1, GSTP1, IL6, MAFK, NAT2, NFKB1, NOS2, NR1I2*, *RIPOR2, SOD1, TGFBRAP1,* and *XPO1*), or pharmacokinetics of isoniazid (*NAT2*) and rifampin (*SLCO1B1*). A list of all genotyped polymorphisms, associated phenotypes, and selected references are provided in **Supplemental Table S1**). Genotyping was done at VANTAGE (Vanderbilt Technology for Advanced Genomics) using MassARRAY® iPLEX Gold (Agena Bioscience™, California, USA) and Taqman (ThermoFisher Scientific, Massachusetts, USA). Assay design is available upon request. We excluded from single variant analyses 2 polymorphisms with minor allele frequencies <5% (rs78872571 in *NFKB1*, rs2070401 in *BACH1*), as well as 4 *NAT2* polymorphisms used to define *NAT2* acetylator status (explained under *<u>Statistical analysis</u>*). Mean genotyping efficiency was 98.8%, and exceeded 95% for all but one polymorphism, which was 93.4%. In this ancestrally diverse cohort, Hardy-Weinberg P-values were not used to exclude polymorphisms, but rather considered in interpreting association results. Analyses ultimately characterized associations with 43 polymorphisms in 20 genes, including 4 polymorphisms which defined *NAT2* acetylator status (**Supplemental Table S1**).

### Ancestry-informative markers

A set of 46 autosomal ancestry-informative markers (AIMs), selected based on a previous analysis of a diverse Brazilian cohort,[27] were genotyped at the Laboratório de Biologia Molecular Aplicada a Micobactérias in Fiocruz, using xMAP multiplexing (Luminex Corporation). These AIMs were used to determine percentage ancestry as described elsewhere [28].

### Whole exome sequencing

A subset of DNA specimens was whole exome sequenced on the Illumina NovaSeq 6000 platform and processed using the DRAGEN Germline application (v.3.8.4) within Illumina’s BaseSpace Sequence Hub (Illumina Inc., San Diego, CA). Gene boundaries were defined using Twist_Exome_plus_RefSeq_GenCode_targets_hg38.bed. The gVCF files were joint-called using the DRAGEN Joint Genotyping Pipeline (v.3.8.4). The joint called VCF file was hard-filtered with a quality score threshold of 10.4139 (error probability <0.01). Quality control of the joint called VCF was done using PLINK2 software [29]. Samples were filtered for call rate (98%) and checked for relatedness and sex assignment. A total of 203,337 bi-allelic single nucleotide variants and short indels were processed, of which 5,772 were novel. This covered 30,466 overlapped genes and 170,658 overlapped transcripts. Variants were annotated using the Ensembl variant effect predictor. To calculate principal components (PCs) using PLINK2, a set of variants present in 1000 Genomes Project phase3 were selected and pruned based on linkage disequilibrium.

### Statistical analysis

*NAT2* genotypes were categorized based on combinations of rs1801280 (*NAT2*5*), rs1799930 (*NAT2*6*), rs1799931 (*NAT2*7*), and rs1801279 (*NAT2*14*): as slow if homozygous for the variant allele at any locus or heterozygous at 2 or more loci; intermediate if heterozygous at a single locus; or rapid if no variant allele [30]. Other individual polymorphisms were analyzed as ordinal variables assuming additive genetic effect. *CYP2B6* metabolizer group was defined based on combinations of three polymorphisms (rs3745274, rs28399499, rs4803419) as described elsewhere [31]. We did not correct for multiple comparisons since only functionally relevant candidate genes or polymorphisms were included. Two-sided tests were used.

Logistic regression analyses assessed associations between outcomes and *NAT2*. To decrease the likelihood of overfitting the models, we used two approaches to adjust for covariates: standard multivariable logistic regression and propensity score-adjusted logistic regression. In fully adjusted models, we adjusted for seven covariates: baseline age, sex, HIV status, hemoglobin A1c (HbA1c) percent, percent African ancestry, percent European ancestry, and prescribed DOT. For outcomes with relatively few events, we used logistic regression with propensity score weights to avoid overfitting [32]. Propensity score models, which are also used as variable reduction techniques in regression models,[33] were used to regress *NAT2* on pre-specified covariates: baseline age, BMI, sex, HIV status, HbA1c percent, prescribed DOT, history of alcohol, tobacco, and drug use. We used multinomial regressions to estimate propensity scores for each patient and outcome. Propensity scores were later used as weights in primary outcome models that regressed outcomes on *NAT2*. Of note, some categories of *NAT2* had very few or no events, a situation denoted as *complete separation*.[34] Standard logistic regression has numerical issues in such settings and cannot be applied. An alternative approach is logistic regression with Firth correction,[35, 36] which reduces the small sample bias of maximum likelihood estimates by maximizing a penalized log-likelihood while also providing a solution to separation.[37] We applied Firth correction to propensity score-weighted logistic regression to obtain estimates for parameters of interest. Since Wald-type confidence intervals may lead to poor coverage with small samples, we used profile likelihood to obtain 95% confidence intervals in Firth-corrected propensity score-weighted logistic regression. Similar analyses were performed using all grade ≥2 and all grade ≥3 ADR, as well as all grade ≥2 hepatic events, regardless of whether they were treatment-related.

We used statistical software R version 4.2.0 [38] and STATA version 17 (StataCorp, College Station, Texas, USA). Associations with the 43 polymorphisms were assessed using PLINK [29].

For whole exome sequencing, single variant and gene-based tests were performed using RVTESTS [39]. Single variant associations were tested on variants with call rates >98%, Hardy-Weinberg equilibrium p >10^-7^, and minor allele count >10. The model was adjusted for age, sex, and 4 PCs. Gene-based tests were performed with the option SKAT-O, which optimizes the generalized SKAT over a grid of values of Rho between 0 and 1, corresponding to a kernel SKAT test and a Burden test. The analysis was run using rare (minor allele frequency <0.05), coding, and loss-of-function variants. Gene boundaries were assigned using the Ensembl GRCh38.p13 genetic map. Tests were adjusted for age, sex, and 4 PCs.

## Results

### Characteristics of the cohort

A total of 963 RePORT-Brazil participants treated for culture-confirmed, drug-susceptible pulmonary tuberculosis were evaluable for ADR and microbiologic outcomes, of whom 937 received standard anti-tuberculosis therapy. After excluding participants in whom genotyping for *NAT2* or AIMs failed, or who lacked baseline data on HIV status, HbA1c, or whether DOT was prescribed, 903 participants were included in genetic association analyses. Derivation of our cohort of 903 participants, stratified by HIV status, is presented in **Figure 1**.

**Figure 1.**
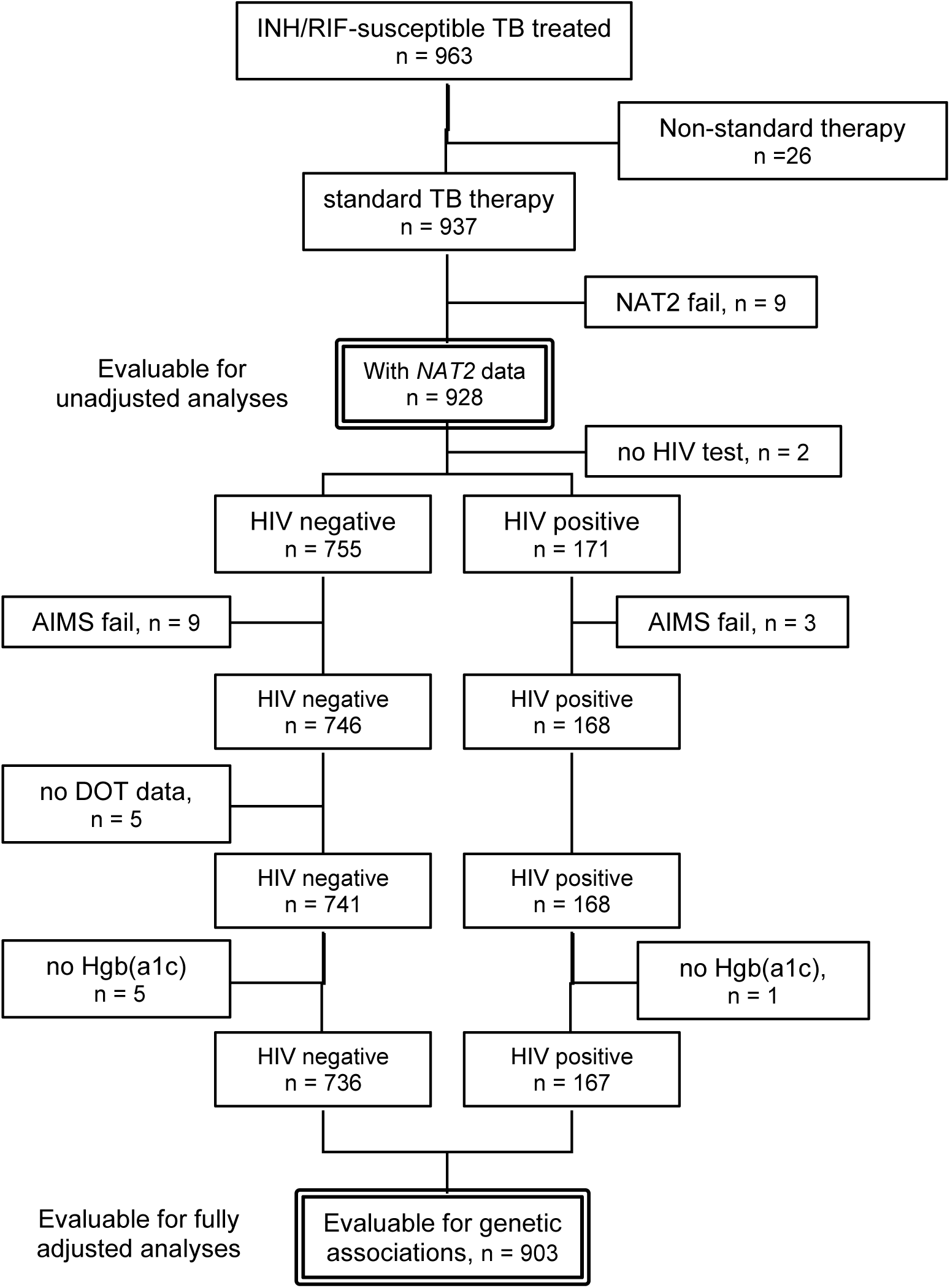
Derivation of the RePORT-Brazil genetic analysis cohort.

Baseline characteristics of participants are presented in **Table 1**. Two-thirds were male, and 167 (18.5%) were PWH. Regarding *NAT2*, there were 77 (8.5%) rapid acetylators, 367 (40.6%) intermediate acetylators, and 459 (50.8%) slow acetylators. Median proportion genetic ancestry based on AIMs was 42% European, 28% African, and 21% Amerindian.

**Table 1.** Baseline characteristics of RePORT-Brazil participants in genetic association analyses.

|  | **Total**<br>**(n=903)** | **HIV-negative**<br>**(n=736)** | **HIV-positive**<br>**(n=167)** | **p-**<br>**value** |
| --- | --- | --- | --- | --- |
| Age in years, median (IQR) | 35.0 (25.0, 48.5) | 35.0 (25.0, 50.0) | 35.0 (27.5, 42.0) | 0.27 |
| Sex, n (%) |  |  |  | 0.004 |
| male | 598 (66.2%) | 471 (64.0%) | 127 (76.0%) |  |
| female | 305 (33.8%) | 265 (36.0%) | 40 (24.0%) |  |
| BMI in kg/m<sup>2</sup>, median (IQR) | 20.2 (18.3, 22.5) | 20.2 (18.3, 22.5) | 19.9 (18.2, 22.1) | 0.58 |
| Directly observed therapy, n (%) | 632 (70.0%) | 506 (68.8%) | 126 (75.4%) | 0.11 |
| HbA1c ≥6.5%, n (%) |  |  |  | 0.66 |
| <5.7 | 229 (25.4%) | 182 (24.7%) | 47 (28.1%) |  |
| 5.7 – 6.4 | 466 (51.6%) | 383 (52.0%) | 83 (49.7%) |  |
| ≥6.5% | 208 (23%) | 171 (23%) | 37 (22.2%) |  |
| NAT2 acetylator; n (%) |  |  |  | 0.68 |
| rapid | 77 (8.5) | 60 (8.2) | 17 (10.2) |  |
| intermediate | 367 (40.6) | 299 (40.6) | 68 (40.7) |  |
| slow | 459 (50.8) | 377 (51.2) | 82 (49.1) |  |
| Alcohol use history, n (%) | 757 (83.8%) | 601 (81.7%) | 156 (93.4%) | <0.001 |
| Tobacco smoking history, n (%) | 470 (52.0%) | 369 (50.1%) | 101 (60.5%) | 0.020 |
| Drug use history, n (%) | 308 (34.1%) | 218 (29.6%) | 90 (53.9%) | <0.001 |
| Ancestry, median (IQR) |  |  |  |  |
| African | 0.28 (0.15, 0.46) | 0.31 (0.17, 0.49) | 0.18 (0.12, 0.28) | <0.001 |
| European | 0.42 (0.28, 0.54) | 0.42 (0.29, 0.54) | 0.41 (0.27, 0.54) | 0.480 |
| Amerindian | 0.21 (0.13, 0.34) | 0.19 (0.13, 0.31) | 0.33 (0.20, 0.51) | <0.001 |

Regarding adverse events, 125 (13.8%) had documented grade ≥2 adverse events during follow-up, including 83 (9.2%) considered tuberculosis treatment related, 49 (5.4%) had grade ≥3 adverse events, including 16 (1.8%) considered tuberculosis treatment related, and 25 (2.8%) had documented grade ≥2 hepatic adverse events. Regarding microbiologic outcomes, 37 (4.1%) experienced treatment failure/recurrence. The distribution of adverse event and microbiologic outcomes, stratified by HIV status, is provided in **Table 2**, and stratified by *NAT2* group in **Supplemental Table S2**.

**Table 2.** Tuberculosis treatment-related adverse drug reactions and treatment outcomes by HIV status.

|  | Total<br>(n=903) | HIV-negative<br>(n=736) | HIV-positive<br>(n=167) | P-value |
| --- | --- | --- | --- | --- |
| Any treatment-related ADR<sup>a</sup> |  |  |  |  |
| Grade $\geq 2$ | 83 (9.2%) | 59 (8.0%) | 24 (14.4%) | 0.02 |
| Grade $\geq 3$ | 16 (1.8%) | 13 (1.8%) | 3 (1.8%) | 1.00 |
| Treatment-related hepatic ADR |  |  |  |  |
| Grade $\geq 2$ | 22 (2.4%) | 14 (1.9%) | 8 (4.8%) | 0.46 |
| Treatment failure or recurrence | 37 (4.1%) | 27 (3.7%) | 10 (6.0%) | 0.25 |

<sup>a</sup> ADR, adverse drug reaction

### NAT2 acetylator status and adverse events

As noted in **Methods**, we used two approaches to assess the effect of *NAT2* on outcomes of interest in a multivariable setting: standard multivariable logistic regression and Firth-corrected propensity score-adjusted models. Results for treatment-related adverse events (grade ≥2, grade ≥3, and hepatoxicity) are shown in **Table 3**. Associations with secondary outcomes (any grade ≥2 and grade ≥3 event) are shown in **Supplemental Table S3**.

**Table 3.**
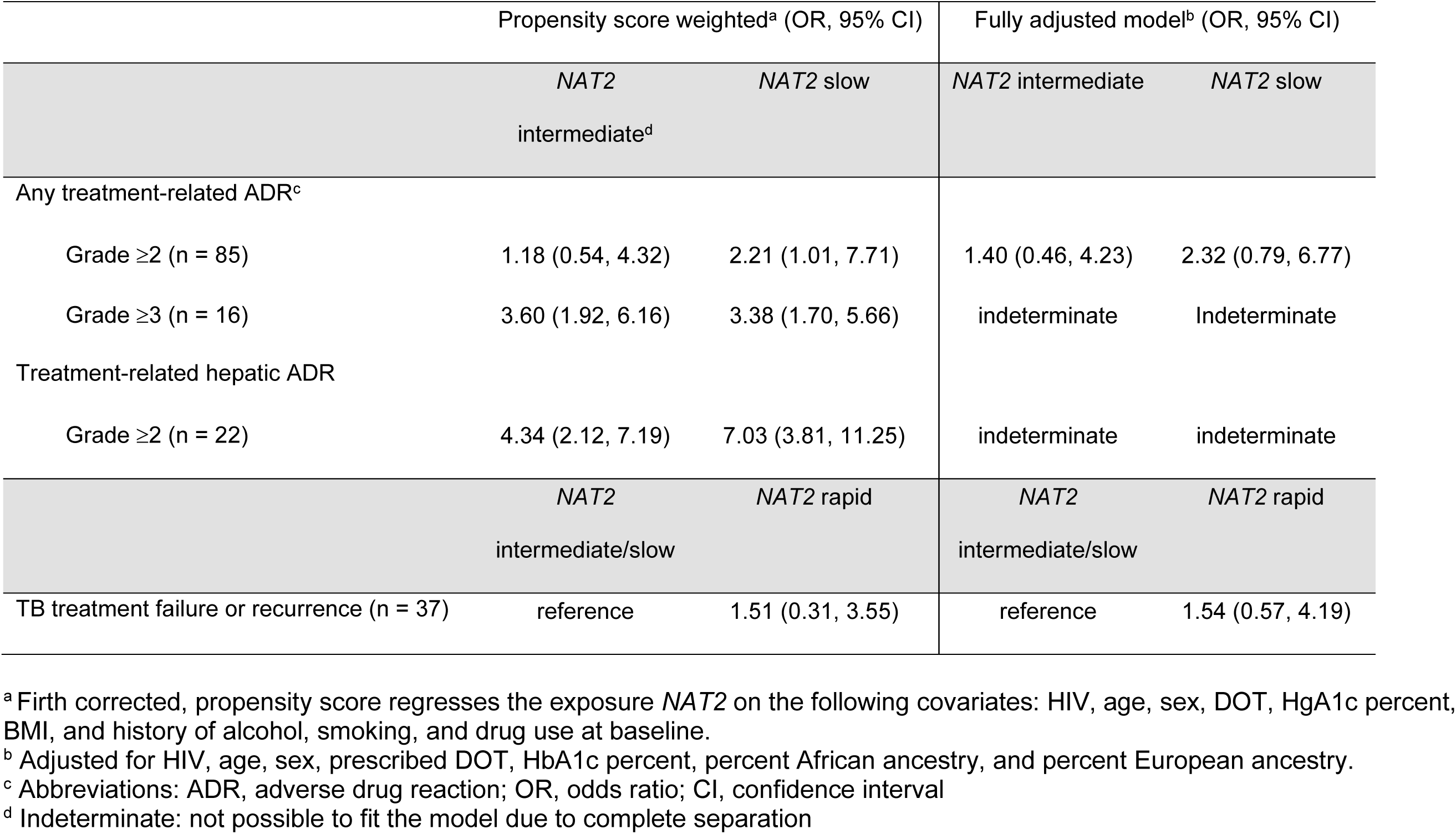
Associations of *NAT2* acetylator status with tuberculosis treatment-related adverse drug reactions and treatment outcome in multivariable models.

Regarding grade ≥2 treatment-related adverse events, both Firth-corrected propensity score-weighted models and standard multivariable logistic regression showed that *NAT2* slow acetylators had higher odds of adverse events compared to rapid acetylators: OR = 2.21 (95% CI = [1.01, 7.71]) and OR = 2.32 (95% CI = [0.79, 6.77]), respectively (**Table 3**). We did not detect differences between intermediate and rapid acetylators. Both approaches had similar point estimates, but different confidence intervals, highlighting differences between Likelihood- profile and Wald-type confidence intervals, and how the latter is affected by finite sample approximations.

Results were similar regarding grade ≥3 treatment-related adverse events. Firth-corrected propensity-score weighted models showed both *NAT2* intermediate and slow acetylators had higher odds of ADR, compared to rapid acetylators: OR = 3.60, 95% CI = [1.92, 6.16] and OR = 3.38, 95% CI = [1.70, 5.66], respectively (**Table 3**). Because there were no events among rapid acetylators (i.e., complete separation; see **Supplemental Table S2**), standard multivariable logistic regression could not be computed.

Regarding treatment-related grade ≥2 hepatic adverse events, Firth-corrected propensity-score weighted models showed both *NAT2* intermediate and slow acetylators had higher odds of ADR, compared to rapid acetylators: OR = 4.34, 95% CI = [2.12, 7.19] and OR = 7.03, 95% CI = [3.81, 11.25], respectively. Standard multivariable logistic regression could not be computed because there were no events in rapid acetylators (**Table 3**).

Similar results were observed when analyzing all ADRs, regardless of whether they were treatment-related. Compared to *NAT2* rapid acetylators, slow acetylators had higher odds of any grade ≥2 ADR, while both intermediate and slow acetylators had higher odds of any grade ≥3 and any grade ≥2 ADR (**Supplemental Table S3**).

### NAT2 acetylator status and treatment failure/recurrence

We hypothesized that risk of treatment failure/recurrence would be increased among *NAT2* rapid acetylators due to lower isoniazid levels. We therefore combined *NAT2* intermediate and slow acetylators as the reference group. In propensity score-weighted models, the odds ratio in rapid acetylators was approximately 1.51 (95% CI = [0.31; 3.55]). Similarly, in fully adjusted models for treatment failure/recurrence, the odds ratio in rapid acetylators was approximately 1.54 (95% CI = [0.57; 4.19]) (**Table 3**).

### Genetic associations after adjusting for NAT2

To determine whether any of the 39 individual polymorphisms (not including the 4 polymorphisms that were used to define *NAT2* acetylator group) in 20 genes were associated with ADR or treatment failure/recurrence outcomes, we performed multivariable logistic regression analyses of each polymorphism while adjusting for *NAT2* acetylator group, baseline age, sex, HIV status, HbA1c percent, percent African ancestry, percent European ancestry, and prescription of DOT. For ADR we treated *NAT2* as an ordinal variable with three levels (rapid, intermediate, and slow), while for failure/recurrence we treated it as two levels (rapid versus intermediate/slow). In these analyses, 4 polymorphisms in 4 genes (*GSTM1, NAT2*, *NR1/2,* and *SLCO1B1*) were associated with at least one treatment-related ADR category at p <0.05 (without correcting for multiple testing), while 3 polymorphisms in 2 genes (*NR1I2* and *SLCO1B1*) and 1 long non-coding RNA (LINC01857) were associated with failure/recurrence. These associations are presented in **Table 4**.

**Table 4.**
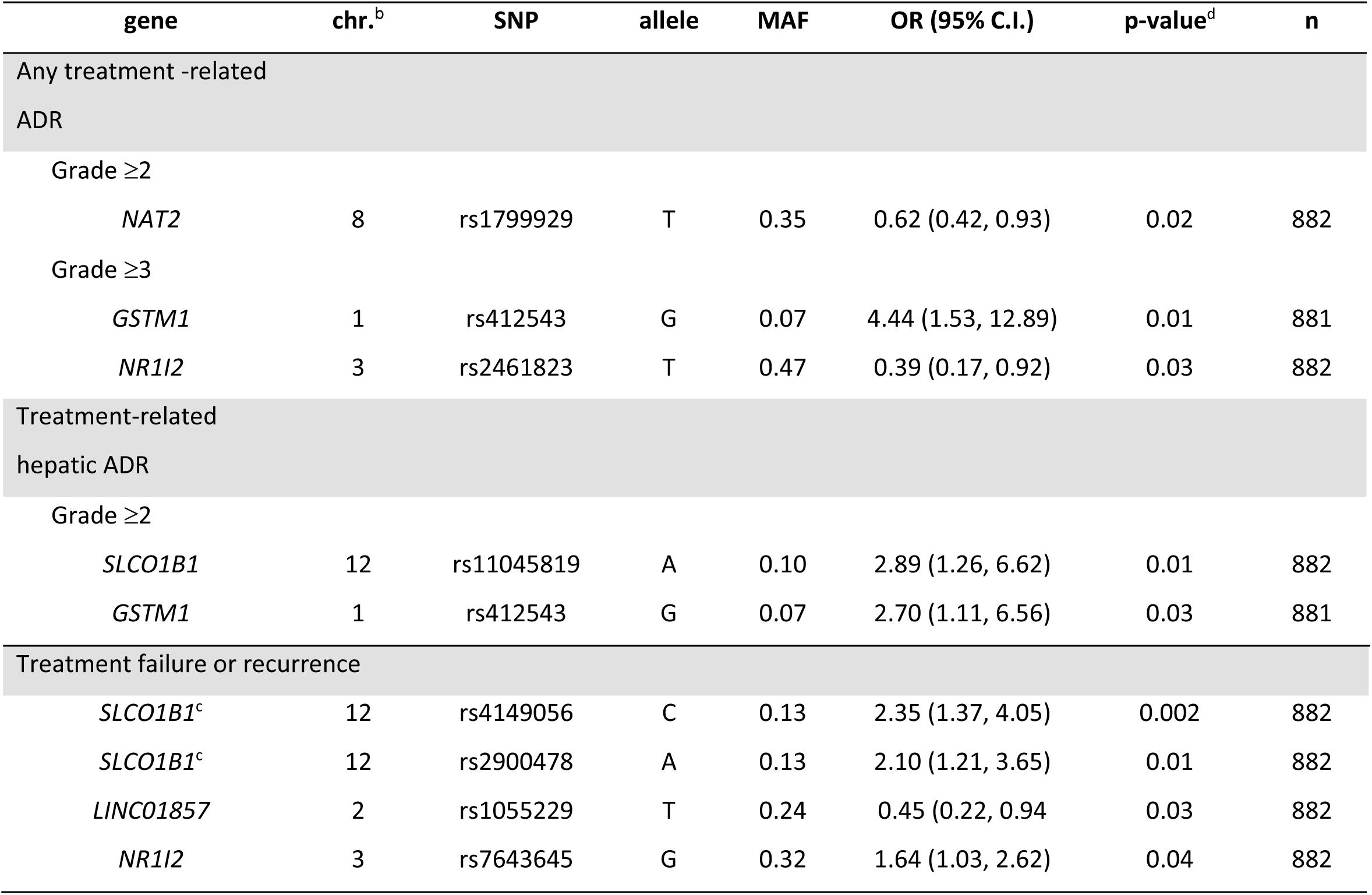
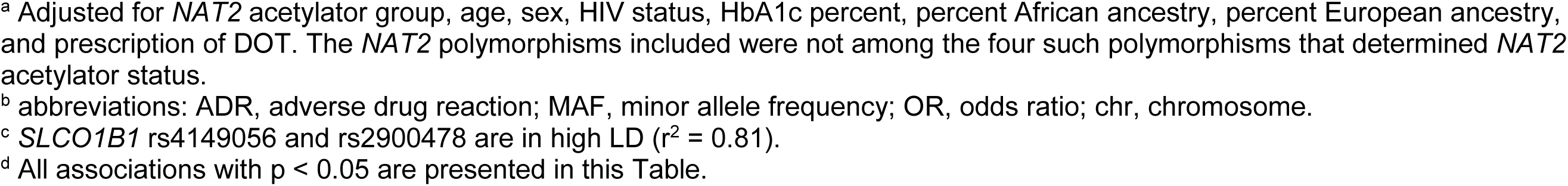
Associations of selected polymorphisms with tuberculosis treatment-related adverse drug reactions and treatment outcome by multivariable models ^a^.

| gene | chr.<sup>b</sup> | SNP | allele | MAF | OR (95% C.I.) | p-value<sup>d</sup> | n |
| --- | --- | --- | --- | --- | --- | --- | --- |
| Any treatment -related ADR |  |  |  |  |  |  |  |
| Grade ≥2 |  |  |  |  |  |  |  |
| *NAT2* | 8 | rs1799929 | T | 0.35 | 0.62 (0.42, 0.93) | 0.02 | 882 |
| Grade ≥3 |  |  |  |  |  |  |  |
| *GSTM1* | 1 | rs412543 | G | 0.07 | 4.44 (1.53, 12.89) | 0.01 | 881 |
| *NR1I2* | 3 | rs2461823 | T | 0.47 | 0.39 (0.17, 0.92) | 0.03 | 882 |
| Treatment-related hepatic ADR |  |  |  |  |  |  |  |
| Grade ≥2 |  |  |  |  |  |  |  |
| *SLCO1B1* | 12 | rs11045819 | A | 0.10 | 2.89 (1.26, 6.62) | 0.01 | 882 |
| *GSTM1* | 1 | rs412543 | G | 0.07 | 2.70 (1.11, 6.56) | 0.03 | 881 |
| Treatment failure or recurrence |  |  |  |  |  |  |  |
| *SLCO1B1*<sup>c</sup> | 12 | rs4149056 | C | 0.13 | 2.35 (1.37, 4.05) | 0.002 | 882 |
| *SLCO1B1*<sup>c</sup> | 12 | rs2900478 | A | 0.13 | 2.10 (1.21, 3.65) | 0.01 | 882 |
| *LINC01857* | 2 | rs1055229 | T | 0.24 | 0.45 (0.22, 0.94) | 0.03 | 882 |
| *NR1I2* | 3 | rs7643645 | G | 0.32 | 1.64 (1.03, 2.62) | 0.04 | 882 |

<sup>a</sup> Adjusted for *NAT2* acetylator group, age, sex, HIV status, HbA1c percent, percent African ancestry, percent European ancestry, and prescription of DOT. The *NAT2* polymorphisms included were not among the four such polymorphisms that determined *NAT2* acetylator status.
<sup>b</sup> abbreviations: ADR, adverse drug reaction; MAF, minor allele frequency; OR, odds ratio; chr, chromosome.
<sup>c</sup> *SLCO1B1* rs4149056 and rs2900478 are in high LD ( $r^2 = 0.81$ ).
<sup>d</sup> All associations with $p < 0.05$ are presented in this Table.

### Whole exome associations

A total of 87 participants (43 cases and 44 controls) were included in whole exome sequence analyses. Cases had grade ≥3 toxicity at least possibly related to tuberculosis treatment, permanent drug discontinuation due to treatment toxicity, or grade ≥2 hepatotoxicity at least possibly related to treatment. One case that failed sex check quality control was excluded. In the single variant analysis, the lowest P-values was rs10991326 in *ORC13C2* (*p* = 2.8 x 10^-5^). The three other lowest P-values were *CPHXL* rs882263 (*p* = 3.2 x 10^-5^), *SULT1C3* rs6772745 (*p* = 4.6 x 10^-5^), and *CHIA* rs2820092 (*p* =8.7 x 10^-5^). The top gene-based result was *WTAPP1* (*p* = 0.0008). In analysis limited to 37 cases with hepatotoxicity and 37 matched controls, the lowest p value was *VTI1A* rs17353359 (p =2.5 x 10^-5^), and the lowest p values for genes were *METTL17* (*p =* 0.0007) and *PRSS57* (*p =* 0.0008). None associations were significant after correcting for multiple testing.

## Discussion

The present study involved 903 adults who received standard therapy for pulmonary tuberculosis during a prospective observational cohort study in Brazil. Compared to *NAT2* rapid acetylators, slow acetylators were at increased risk for grade ≥2 treatment-related ADR, while both slow and intermediate acetylators were at increased risk for grade ≥3 treatment-related ADR, and for grade ≥2 treatment-related hepatic ADR. The association of *NAT2* acetylator status with risk of hepatotoxicity during tuberculosis treatment was expected. Prior meta-analyses showed increased risk among slow acetylators [21–23, 40]. The largest such analysis, which included 38 studies with 2,225 patients and 4,906 controls, reported an odds ratio of 3.2 (95% CI: 2.5–4.1)[23]. In the present study, rapid acetylators had a higher odds ratio for failure/recurrence, but this was not statistically significant, possible due to the few events.

We determined *NAT2* acetylator status based on four signature polymorphisms that identify *NAT2 *5*, *\*6*, *\*7* and *\*14* loss-of-function alleles. We considered including rs1041983 based on a previous report suggesting its added utility in inferring slow acetylator alleles [41]. However, in that report, a four-polymorphism panel of rs1041983 plus *\*5*, *\*6*, and *\*7* signature polymorphisms performed no better than the three signature polymorphisms. Only with addition of the *\*14* polymorphism did performance improve [41]. In the present study, *NAT2* polymorphisms that were not used to determine acetylator status (rs1041983, rs1208, and rs1799929) were analyzed individually in models that controlled for acetylator status.

In our cohort, standard tuberculosis therapy was generally effective and well-tolerated. Among the 903 participants, 4.1% had treatment failure/recurrence, 5.4% had grade ≥3 ADR (of which 32% were treatment-related), and 2.8% had grade ≥2 hepatic ADR (of which 88% were treatment-related). A recent study from Brazil reported that at least 11% of participants had a grade ≥3 ADR and 9% had liver and biliary system disorders.[42] A previous retrospective study in Brazil reported that 5-20% of individuals receiving standard tuberculosis therapy experienced minor/mild ADR, while 5% on isoniazid, rifampicin and pyrazinamide had hepatic transaminase elevations,[43] consistent with our study. In a recent tuberculosis treatment shorting trial, the standard treatment arm was associated with 1% failure, <1% recurrence, and 1% changed treatment due to ADRs, although nearly 20% experienced grade ≥3 ADRs, of which 3% were hepatic.[4]

In analyses that controlled for *NAT2* acetylator status, as well as clinical covariates, eight additional polymorphisms in *NAT2*, *GSTM1*, *NR1I2*, *SLCO1B1*, as well as in a long non-coding RNAs (lncRNA), were associated with treatment-related ADR or failure/recurrence at P <0.05. Regarding ADR, the *NR1I2* rs2461823 A allele, with a MAF of 47%, was associated with increased grade ≥3 ADR, which is generally consistent with a previous report that associated this allele with increased severity of nonalcoholic fatty liver disease (NAFLD) [44]. The *NAT2* rs1799929 T allele, a tagging polymorphism with a MAF of 35%, was associated with decreased grade ≥2 ADR, but has not been previously associated with tuberculosis treatment outcomes. Similarly, the *SLCO1B1* rs11045819 A allele, a tagging polymorphism with a MAF of 10%, was associated with increased grade ≥2 ADR, has not been previously associated with tuberculosis treatment outcomes. The *GSTM1* rs412543 G allele, with a MAF of 7%, was associated with both grade ≥3 ADR and grade ≥2 hepatic ADR.

Regarding treatment failure/recurrence, the lncRNA (*LINC01875* rs1055229) T allele, with a MAF of 24%, was associated with decreased treatment failure/recurrence, but a previous study associated this allele with increased tuberculosis drug-induced hepatotoxicity [45]. The *NR1I2* rs7643645 G allele was associated with treatment failure/recurrence, but a previous report associated this with increased NAFLD severity [44]. The *SLCO1B1* rs4149056 C allele, with a MAF of 13%, was associated with increased failure/recurrence (as was the linked *SLCO1B1* rs2900478). While rs4149056 C allele increases plasma exposure of organic anion transporting polypeptide 1B1 substrates (e.g., statins), it is not known to affect anti-tuberculosis medications such as rifampin.

Whole exome sequencing provided interesting findings of nominal statistical significance. In single variant analyses, *SULT1C3* rs6772745 was a top hit in both primary and secondary analyses. *SULT1C3* encodes sulfotransferase 1C3, which has activity towards various substrates including hormones, neurotransmitters, drugs, and xenobiotics [46], although its mRNA has been detected only in human intestine, not in liver [47]. *CPHXL* rs882263 was the only top result with an effect in the negative direction. *CPHXL* is predicted to regulate transcription by RNA polymerase II. The top gene-based result, *WTAPP1*, is a pseudogene reported to affect cancer cell behavior and disease progression [48, 49].

This study had limitations. Only 1.8% of participants had grade ≥3 ADR, 2.4% had grade ≥2 hepatic ADR, and 4.1% had failure/recurrence. More cases would have increased power to detect associations. For most participants, we only had aggregated monthly DOT data, so could not reliably assess the impact of adherence. Because participants were receiving multiple medications, and we cannot attribute causation to specific medications.

In summary, *NAT2* slow acetylator status was associated with increased risk for tuberculosis treatment-related ADR, including hepatic ADR, in a clinical cohort representing diverse ancestries from several regions of Brazil. The odds ratio for treatment failure/recurrence was higher among rapid acetylators. Polymorphisms in other genes (*GSTM1*, *NR1I2*, *SLCO1B1*) and in a lncRNA were independently associated with ADR and failure/recurrence. This study extends our understanding of the pharmacogenetics of tuberculosis therapy, and highlights the need for studies in larger cohorts. Studies should evaluate the effect drug dosing informed by host genetics on treatment outcomes.

## Data Availability

All data produced in the present study are available upon reasonable request to the authors

## Acknowledgments

We are grateful to Lauren S. Peetluk, Brian C. Hachey, and Leticia Linhares at Vanderbilt University Medical Center, and Edward P. Acosta at University of Alabama at Birmingham, for their work in support of R01 AI120790.

## Notes

Funding: This work was supported by the Departamento de Ciência e Tecnologia–Secretaria de Ciência e Tecnologia–Ministério da Saúde, Brazil (25029.000507/2013-07); National Institute of Allergy and Infectious Diseases (NIAID) (U01 AI069923, R01 AI120790, U01 AI172064, R01 AI077505, P30 AI110527) and the National Center for Advancing Translational Science (UL1 TR000445) at the National Institutes of Health.

### Competing Interest Statement

The authors have declared no competing interest.

### Funding Statement

This work was supported by the Departamento de Ciência e Tecnologia, Secretaria de Ciência e Tecnologia, Ministério da Saúde, Brazil (25029.000507/2013-07); National Institute of Allergy and Infectious Diseases (NIAID) (U01 AI069923, R01 AI120790, U01 AI172064, R01 AI077505, P30 AI110527) and the National Center for Advancing Translational Science (UL1 TR000445) at the National Institutes of Health.

### Author Declarations

All clinical investigations were conducted according to the principles of the Declaration of Helsinki. The RePORT-Brazil protocol, informed consent, and study documents were approved by the institutional review boards at each study site and at Vanderbilt University Medical Center. Participation in RePORT-Brazil was voluntary, and written informed consent was obtained from all such participants. 1) Vanderbilt University Medical Center: a. Name of Ethics Committee: Human Research Protections Program; Health Sciences Committee 3 IRB 2) Instituto Nacional de Infectologia, FIOCRUZ: a. Name of Ethics Committee: Instituto Nacional de Infectologia Evandro Chagas; INI /FIOCRUZ IRB 3) Centro Municipal de Saúde (CMS) de Duque de Caxias: a. Name of Ethics Committee: Universidade do Grande Rio Professor José de Souza Herdy; UNIGRANRIO IRB 4) Instituto Brasileiro para Investigação da Tuberculose (IBIT): a. Name of Ethics Committee: Maternidade Climério de Oliveira; UFBA IRB 5) Fundação de Medicina Tropical (FMT): a. Name of Ethics Committee: Fundação de Medicina Tropical; Doutor Heitor Vieira Dourado, IRB

