## Supplemental Tables S1,S2, & S3 for "Pharmacogenetics of tuberculosis treatment toxicity and effectiveness in a large Brazilian cohort"

**Supplemental Table S1: Summary of polymorphisms included in analyses**

This Table includes all 45 polymorphisms in 10 genes relevant to anti-tuberculosis drug-induced hepatotoxicity or anti-tuberculosis drug pharmacokinetics. These include 4 polymorphisms which were used to define *NAT2* acetylator status, and 2 polymorphisms which were excluded from association analyses due to minor allele frequencies <5%. In addition, this Table includes 3 polymorphisms which were used to define *CYP2B6* metabolizer group.

| rs number | chr. | coordinate position[a] | gene | Allele 1 | Allele 2 | minor allele frequency | phenotype | Reference |
|---|---|---|---|---|---|---|---|---|
| rs393994 | 1 | 48643073 | *AGBL4* | G | A | 0.4434 | ATDH | [1] |
| rs319952 | 1 | 48647950 | *AGBL4* | G | A | 0.4658 | ATDH | [1] |
| rs320003 | 1 | 48661106 | *AGBL4* | A | G | 0.4608 | ATDH | [1] |
| rs412543 | 1 | 109687322 | *GSTM1* | G | C | 0.06726 | Drug toxicity | [2] |
| rs11125883 | 2 | 61483438 | *XPO1* | C | A | 0.2632 | ATDH | [3] |
| rs17687727 | 2 | 105249510 | *TGFBRAP1* | A | G | 0.2189 | ATDH | [4] |
| rs1055229 | 2 | 207666959 | *LINC01857* | T | C | 0.2447 | ATDH | [5] |
| rs3814055 | 3 | 119781188 | *NR1I2* | T | C | 0.3046 | ATDH | [6] |
| rs2461823 | 3 | 119801278 | *NR1I2* | T | C | 0.4737 | ATDH | [7] |
| rs7643645 | 3 | 119806650 | *NR1I2* | G | A | 0.3208 | ATDH | [7] |
| rs78872571 | 4 | 102512308 | *NFKB1* | T | C | 0.0213 | ATDH | [8] |
| rs4647992 | 4 | 102534190 | *NFKB1* | T | C | 0.05767 | ATDH | [8] |
| rs10946737 | 6 | 24967012 | *RIPOR2* | A | G | 0.2301 | ATDH | [1, 9] |
| rs10946739 | 6 | 24992899 | *RIPOR2* | T | C | 0.2217 | ATDH | [1] |
| rs4720833 | 7 | 1534767 | *MAFK* | A | G | 0.3242 | ATDH | [10] |
| rs1800796 | 7 | 22726627 | *IL6* | C | G | 0.1327 | ATDH | [11] |
| rs1045642 | 7 | 87509329 | *ABCB1* | A | G | 0.3813 | ATDH | [12] |
| rs4646244 | 8 | 18390208 | *NAT2* | A | T | 0.2207 | NAT2 alleles | [13] |
| rs1801279 | 8 | 18400194 | *NAT2* | A | G | 0.03303 | INH PK | [13] |
| rs1041983 | 8 | 18400285 | *NAT2* | T | C | 0.3711 | NAT2 alleles | [13] |
| rs1801280 | 8 | 18400344 | *NAT2* | C | T | 0.3698 | INH PK | [13] |
| rs1799929 | 8 | 18400484 | *NAT2* | T | C | 0.3488 | NAT2 alleles | [13] |

| rs number | chr. | coordinate position[a] | gene | Allele 1 | Allele 2 | minor allele frequency | phenotype | Reference |
|---|---|---|---|---|---|---|---|---|
| rs1799930 | 8 | 18400593 | *NAT2* | A | G | 0.2234 | INH PK | [13] |
| rs1208 | 8 | 18400806 | *NAT2* | G | A | 0.4099 | NAT2 alleles | [13] |
| rs1799931 | 8 | 18400860 | *NAT2* | A | G | 0.08063 | INH PK | [13] |
| rs1495741 | 8 | 18415371 | *NAT2* | G | A | 0.3369 | NAT2 alleles | [13] |
| rs1457043 | 8 | 58497880 | *CYP7A1* | C | T | 0.4239 | ATDH | [14] |
| rs2273697 | 10 | 99804058 | *ABCC2* | A | G | 0.1738 | ATDH | [15] |
| rs3740065 | 10 | 99845936 | *ABCC2* | G | A | 0.1573 | ATDH | [16] |
| rs2031920 | 10 | 133526341 | *CYP2E1* | T | C | 0.07584 | ATDH | [17] |
| rs1695 | 11 | 67585218 | *GSTP1* | G | A | 0.3992 | ATDH | [18] |
| rs4149032 | 12 | 21164857 | *SLCO1B1* | T | C | 0.4803 | RIF PK | [19] |
| rs4149034 | 12 | 21164988 | *SLCO1B1* | A | G | 0.463 | Tag SNP | [19] |
| rs2417957 | 12 | 21170677 | *SLCO1B1* | T | C | 0.1406 | Tag SNP | [19] |
| rs2306283 | 12 | 21176804 | *SLCO1B1* | A | G | 0.4022 | Tag SNP | [19] |
| rs11045819 | 12 | 21176879 | *SLCO1B1* | A | C | 0.1025 | Tag SNP | [19] |
| rs4149056 | 12 | 21178615 | *SLCO1B1* | C | T | 0.1299 | SLCO1B1 activity, statin tox. | [19] |
| rs1564370 | 12 | 21182256 | *SLCO1B1* | G | C | 0.3102 | Tag SNP | [19] |
| rs4149063 | 12 | 21197856 | *SLCO1B1* | T | G | 0.1943 | Tag SNP | [19] |
| rs2900478 | 12 | 21215863 | *SLCO1B1* | A | T | 0.1349 | Tag SNP | [19] |
| rs4842407 | 12 | 78807293 | *lincRNA* | G | A | 0.421 | ATDH | [20] |
| rs7958375 | 12 | 111202213 | *CUX2* | A | G | 0.05991 | ATDH | [1] |
| rs11080344 | 17 | 27777485 | *NOS2* | T | C | 0.3729 | ATDH | [3] |
| rs4803419 | 19 | 41006887 | *CYP2B6* | T | C | 0.2805 | EFV PK | [21] |
| rs3745274 | 19 | 41006936 | *CYP2B6* | T | G | 0.3678 | EFV PK | [21] |
| rs28399499 | 19 | 41012316 | *CYP2B6* | C | T | 0.02856 | EFV PK | [21] |
| rs2070401 | 21 | 29343164 | *BACH1* | G | A | 0.03863 | ATDH | [3] |
| rs2070424 | 21 | 31667007 | *SOD1* | G | A | 0.1803 | ATDH | [22] |

[a] Based on genome build GRCh38

Selected References

**Supplemental Table S2. Frequency of adverse drug reactions and treatment outcomes by NAT2 acetylator group.**

| | **Total (n = 903)** | **Rapid acetylator (n = 77)** | **Intermediate acetylator (n = 367)** | **Slow acetylator (n = 459)** |
|---|---|---|---|---|
| Any adverse drug reaction | | | | |
| Grade ≥2 | 125 (13.8%) | 6 (7.8%) | 44 (12.0%) | 75 (16.3%) |
| Grade ≥2 TB treatment-related | 83 (9.2%) | 4 (5.2%) | 27 (7.4%) | 52 (11.3%) |
| Grade ≥3 | 49 (5.4%) | 1 (1.3%) | 22 (6.0%) | 26 (5.7%) |
| Grade ≥3 TB treatment-related | 16 (1.8%) | 0 (0.0%) | 8 (2.2%) | 8 (1.7%) |
| Hepatic adverse drug reaction | | | | |
| Grade ≥2 hepatic | 25 (2.8%) | 0 (0.0%) | 9 (2.5%) | 16 (3.5%) |
| Grade ≥2 TB treatment-related hepatic | 22 (2.4%) | 0 (0.0%) | 8 (2.2%) | 14 (3.1%) |
| Treatment failure or recurrence | 37 (4.1%) | 5 (6.5%) | 13 (3.5%) | 19 (4.1%) |

**Supplemental Table S3. Associations of *NAT2* genotype with any adverse drug reaction.**

| | Propensity score weighted[a] (OR, 95% CI) | | Fully adjusted model[b] (OR, 95% CI) | |
|---|---|---|---|---|
| | *NAT2* intermediate (n = 367) | *NAT2* slow (n = 459) | *NAT2* intermediate (n = 367) | *NAT2* slow (n = 459) |
| Any adverse drug reaction[c] | | | | |
| Grade ≥2 (n = 125) | 1.40 (0.67, 4.12) | 2.25 (1.1, 6.48) | 1.66 (0.70, 4.63) | 2.45 (1.06, 6.69) |
| Grade ≥3 (n = 49) | 3.23 (1.06, 13.58) | 3.37 (1.18, 13.44) | 4.95 (1.00, 89.88) | 5.05 (1.03, 91.37) |
| Hepatic adverse drug reaction[c] | | | | |
| Grade ≥2 (n = 25) | 4.34 (2.12, 7.19) | 7.03 (3.81, 11.25) | indeterminate | indeterminate |

[a] Firth corrected, propensity score regresses the exposure *NAT2* on the following covariates: HIV, age, sex, DOT, HbA1c percent, BMI, and history of alcohol, smoking, and drug use at baseline.
[b] Profile-likelihood confidence intervals
[b] Adjusted for HIV, age, sex, DOT, HbA1c percent, percent African ancestry, percent European ancestry.
[b] Wald-type confidence intervals
[c] Reference level: *NAT2* rapid
Abbreviations: OR, odds ratio; CI, confidence interval
Indeterminate: not possible to fit the model due to complete separation.
